# All-cause excess mortality in the State of Gujarat, India, during the COVID-19 pandemic (March 2020-April 2021)

**DOI:** 10.1101/2021.08.22.21262432

**Authors:** Rolando J Acosta, Biraj Patnaik, Caroline Buckee, Mathew V Kiang, Rafael A Irizarry, Satchit Balsari, Ayesha S Mahmud

**Affiliations:** Department of Biostatistics, Harvard T. H. Chan School of Public Health, Boston, MA 02115; National Foundation for India, New Delhi 110 003, India; Center for Communicable Disease Dynamics, Harvard T. H. Chan School of Public Health, Boston, MA 02115; Department of Epidemiology, Harvard T. H. Chan School of Public Health, Boston, MA 02115; Department of Epidemiology and Population Health, Stanford University School of Medicine, California USA; Department of Data Science, Dana-Farber Cancer Institute, Boston, MA, USA; Department of Emergency Medicine, Beth Israel Deaconess Medical Center, Harvard Medical School, Boston, MA 02115, USA; Department of Global Health and Population, Harvard T. H. Chan School of Public Health, Boston, MA 02115; Department of Demography, University of California, Berkeley, CA 94720

## Abstract

Official COVID-19 mortality statistics are strongly influenced by local diagnostic capacity, strength of the healthcare and vital registration systems, and death certification criteria and capacity, often resulting in significant undercounting of COVID-19 attributable deaths. Excess mortality, which is defined as the increase in observed death counts compared to a baseline expectation, provides an alternate measure of the mortality shock – both direct and indirect – of the COVID-19 pandemic. Here, we use data from civil death registers from a convenience sample of 90 municipalities across the state of Gujarat, India, to estimate the impact of the COVID-19 pandemic on all-cause mortality. Using a model fit to weekly data from January 2019 to February 2020, we estimated excess mortality over the course of the pandemic from March 2020 to April 2021. We estimated 21,300 [95% CI: 20,700, 22,000] excess deaths across these municipalities in this period, representing a 44% [95% CI: 43%, 45%] increase over the expected baseline. The sharpest increase in deaths in our sample was observed in late April 2021, with an estimated 678% [95% CI: 649%, 707%] increase in mortality from expected counts. The 40 to 65 age group experienced the highest increase in mortality relative to the other age groups. We found substantial, yet similar, increases in mortality for males and females. Our excess mortality estimate for these 90 municipalities, representing approximately 5% of the state’s population, exceeds the official COVID-19 death count for the entire state of Gujarat, even before the delta wave of the pandemic in India peaked in May 2021. Prior studies have concluded that true pandemic-related mortality in India greatly exceeds official counts. This study, using data directly from the first point of official death registration data recording, provides incontrovertible evidence of the high excess mortality in Gujarat from March 2020 to April 2021.

## 1 Introduction

Official COVID-19 mortality counts around the world largely rely on the attribution of COVID-19 as a cause of death on death certificates[1, 2, 3]. Data from death certificates are then aggregated to centralized databases, often with some reporting delay[4, 5, 6]. These data can be assumed to be sensitive and reliable indicators of the true national toll of COVID-19 related deaths when most patients with COVID-19 have access to healthcare, health care providers have the knowledge and tools necessary to diagnose COVID-19, and those recording deaths on death certificates have the requisite training to note COVID-19 as an underlying cause of death on the certificate[2, 7]. One or more of these conditions is often not met, resulting in underestimates of COVID-19 related deaths, as is the case in India[8, 9].

In the absence of reliable death registration data in the aftermath of disasters and public health emergencies, scientists have relied on alternative methods to estimate deaths, including household based surveys, crematoria and funeral home body counts, or verbal autopsies[10, 11, 12, 13]. In many countries, the estimation of all-cause mortality has provided an alternative proxy for the underestimation of COVID-19 attributable deaths in official statistics, whereby total observed deaths are compared to expected deaths computed from historical baselines[14, 15, 16, 17, 18, 19]. During disasters and public health crises, all-cause mortality estimates can provide an overall measure of the mortality shock. During the COVID-19 pandemic, all-cause mortality data include both directly attributable deaths (those that died from SARS-Cov-2 and its complications) and indirectly attributable deaths (those that died from the indirect impacts of the pandemic, like disruptions in access to livelihoods, food security, public assistance, preventive health interventions, or medical care)[20, 21].

Few studies have looked at excess mortality calculations in resource poor settings where baseline data are not readily available. Studies, news reports, and lived experience suggests that the true pandemic-related death toll in India is larger than the official estimate[22, 23]. Here, we use deaths data from multiple municipalities in the State of Gujarat in India to examine the impact of the COVID-19 pandemic of all-cause mortality; we explore differences in death counts by age and sex over the course of the pandemic, and estimate excess mortality resulting from the pandemic.

### 2 Methods

We used de-identified data from civil death registers, the official records at the municipal jurisdictional level. The data comprise a convenience sample of 92 municipalities, made publicly available by The Reporter’s Collective (RC), a group of investigative journalists in India[24, 25]. At the time of our analysis, RC had collected and made public, data from January 2020 to April 2021. Deaths are first recorded in the death registers maintained at the Gram Panchayats (village committees) in rural India, and in municipalities and municipal corporations in urban India, typically within a few weeks of the death. The data are then aggregated at the district level and submitted to the national Civil Registration System (CRS)[26, 27]. It may take up to nine months from the end of the financial year for the CRS to be fully updated and validated by the government of India. Therefore, data collated directly from the death registers and maintained locally are the most comprehensive source of deaths from 2020 and 2021 currently available - until the CRS is fully updated.

We conducted a secondary analysis of these data directly derived from death registers from 92 (of 162) municipalities in the state of Gujarat, representing a population of at least 3.2 million (according to the 2011 census), or approximately 5% of the total state population of 69 million[28]. These data encompass all recorded deaths from January 2019 to April 2021, and include date of death, date of registration, gender, age, and place of death information. As publicly available data were used to conduct this secondary analysis, the study was IRB exempt.

In Gujarat, according to the National Family Health Survey (NFHS) 2019-20, 93% of all deaths “of usual residents of households” in Gujarat are recorded in the civil death registers. Death registration completeness reaches nearly 96% in urban areas, 92% in rural areas, 94% for males, and 91% for females, overall[29]. Using these records, we computed weekly mortality counts for males and females, and by four age groups: less than 20, 20 to 40, 40 to 65, and 65 years and over, from January 2019 to April 2021.

### 2.1 Statistical Analysis

Based on the reported COVID-19 case and mortality data in India from early 2020, we considered data from January 2019, to February 2020 to represent baseline mortality. We compared the baseline mortality data to observed counts from March 2020 onwards to estimate excess mortality. Due to the paucity of data, ascertaining excess mortality by both age and sex together is not possible. As an alternative, we conducted three analyses. In the first, we aggregated data across all demographic indicators for each municipality, thus we had weekly death counts per jurisdiction. For the second and third analyses, we aggregated data by gender and the age groups mentioned above, respectively. We refer to the three analyses as the *overall* analysis, the *gender* analysis, and the *age* analysis.

To estimate expected mortality over the course of the pandemic we, assume that *Y*_*tmd*_∼ Poisson (*μ*_*tmd*_), where *Y*_*tmd*_is the number of deaths at week *t* in municipality *m* for the demographic group *d*. Note that the index *d* is ignored in the marginal analysis. The mean of the distribution is modeled as

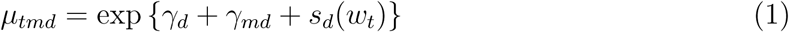

for *t* = 1, …, *T, m* = 1, …, *M*, and *d* = 1, …, *D*. In model 1, *μ*_*tmd*_ represents the average number of deaths at week *t* in municipality *m* for demographic group *d, γ*_*d*_ is a demographic-specific intercept that represents the average number of deaths per week across all municipalities, *γ*_*md*_ is a municipality-specific intercept that represent deviations in the average number of deaths per week in municipality *m* from *γ*_*d*_, *s* is a harmonic component that accounts for seasonality, *w*_*t*_∈ {1, …, 52} corresponds to the week of the year associated with *t, T* is the number of observations in the time-series, *M* is the number of municipalities, and *D* is the number of demographic groups. As a consequence of unreliable population estimates, we assumed that the population size in each municipality was constant from 2019 to 2021, and hence we did not include a population offset in our mean model. We used data from January 2019 to February 2020 to fit model 1 via maximum likelihood assuming an overdispersed Poisson distribution. Then, we estimated the expected mortality for each municipality from March 2020 to April 2021 using the estimated model parameters. To account for variation in the observed counts during the period of interest, let *t*^′^ be a week after the onset of Covid-19 with corresponding outcome *Y*_*t*_*′*_*md*_∼ Poisson (*μ*_*t*_*′*_*md*_). For each municipality, we fit the following model:

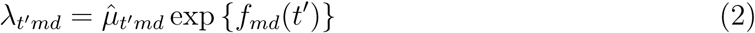

with *λ*_*t*_*′*_*md*_the average number of deaths at *t*^′^, *f*_*md*_a smooth function of time that represents deviations from expected mortality based on historical data, and 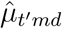 an offset representing expected mortality at *t*^′^. We modeled *f*_*md*_with a natural cubic spline where the number of internal nodes depended on data quality. As before, we fit model 2 via maximum likelihood assuming an overdispersed Poisson distribution. We estimated excess deaths at *t*^′^ with:

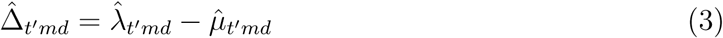

with variance

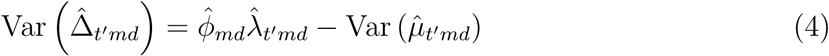

where 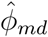 is the estimated dispersion parameter from model 2. We estimated cumulative excess death and associated confidence intervals by summing the excess death estimates and corresponding variance estimates, respectively. Finally, we computed municipality-specific percent changes from average mortality as a function of time, and amalgamated these metrics to ascertain deviations in mortality from expectation for all municipalities combined[19]. In the Results section, we round our excess death estimates proportional to the standard error. For example, if the standard error is in the hundreds, we round our estimate to the nearest tens.

For all three analyses, we removed the municipalities of Jetpur and Modasa due to poor data quality (Supplemental Figure 1). For the gender analysis, we only considered the male and female categories (due to coding inconsistencies across municipalities), and further removed the municipalities of Boriavi, Dhoraji, Halol, Halvad, Idar, Kathlal, Okha, Pardi, and Zalod due to poor data quality (Supplemental Figure 2). Finally, for the age analysis, we categorized the data into the four age groups: less than 20 years, 20 to 40, 40 to 65, and 65 and over. We removed Bharuch, Idar, and Jasdan, also due to poor data quality. For the two youngest groups, we further removed more municipalities for the same reason (Supplemental Table 1). The data and code to reproduce our analysis is available at: http://github.com/RJNunez/gujarat-ed-study.

## 3 Results

### 3.1 Excess deaths overall

77,781 total deaths were recorded across the 90 municipalities over the course of the pandemic, from March 2020 onwards. While deaths were higher in both 2020 (50,866) and 2021 (26,915 up to April) compared to 2019 (42,246), the sharpest increase in deaths was observed during the period encompassing the start of the second wave of the pandemic in March and April 2021 (Figure 1). From our overall analysis, we estimated 21,300 excess deaths [95% CI: 20,700, 22,000] across the 90 municipalities since March 2020, most of which occurred at the start of the second wave in March 2021 (Supplemental Figure 3).

**Figure 1:**
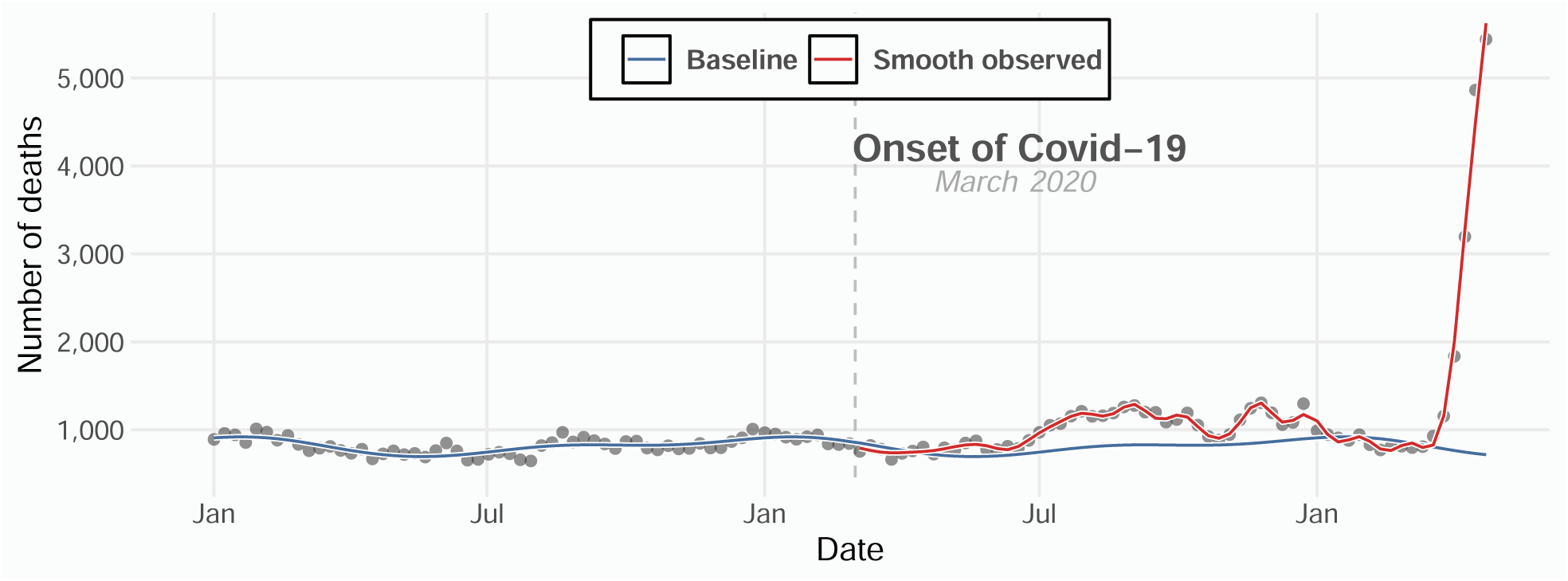
Model fit for weekly death counts amalgamated from multiple municipalities in Gujarat, India. The gray data points are weekly death counts, the dashed-vertical line represents the onset of Covid-19, the blue curve represents the expected weekly death counts based on historical data, and the red curve represents the smooth observed weekly death counts during Covid-19.

### 3.2 Percent change in mortality overall, by age, and by sex

During the first wave of the pandemic, all-cause mortality started to increase in May 2020 and continued to exceed the expected baseline until January 2021 (Figure 2). Over this time period, we estimated a 30.5% [95% CI: 29.4%, 31.6%] increase in mortality from our overall analysis, a 32.2% [95% CI: 30.9%, 33.6%] and 28.2% [95% CI: 26.7%, 29.8%] increase for males and females, respectively, and a 33.6% [95% CI: 31.9%, 35.3%] and 34% [95% CI: 32.5%, 35.5%] increase for the 40 to 65 and the 65 years and over cohort, respectively. The 20 to 40 years age group experienced a 4% [95% CI: 1.91%, 6.09%] increase and we found no evidence of excess mortality for the youngest group during the first wave.

**Figure 2:**
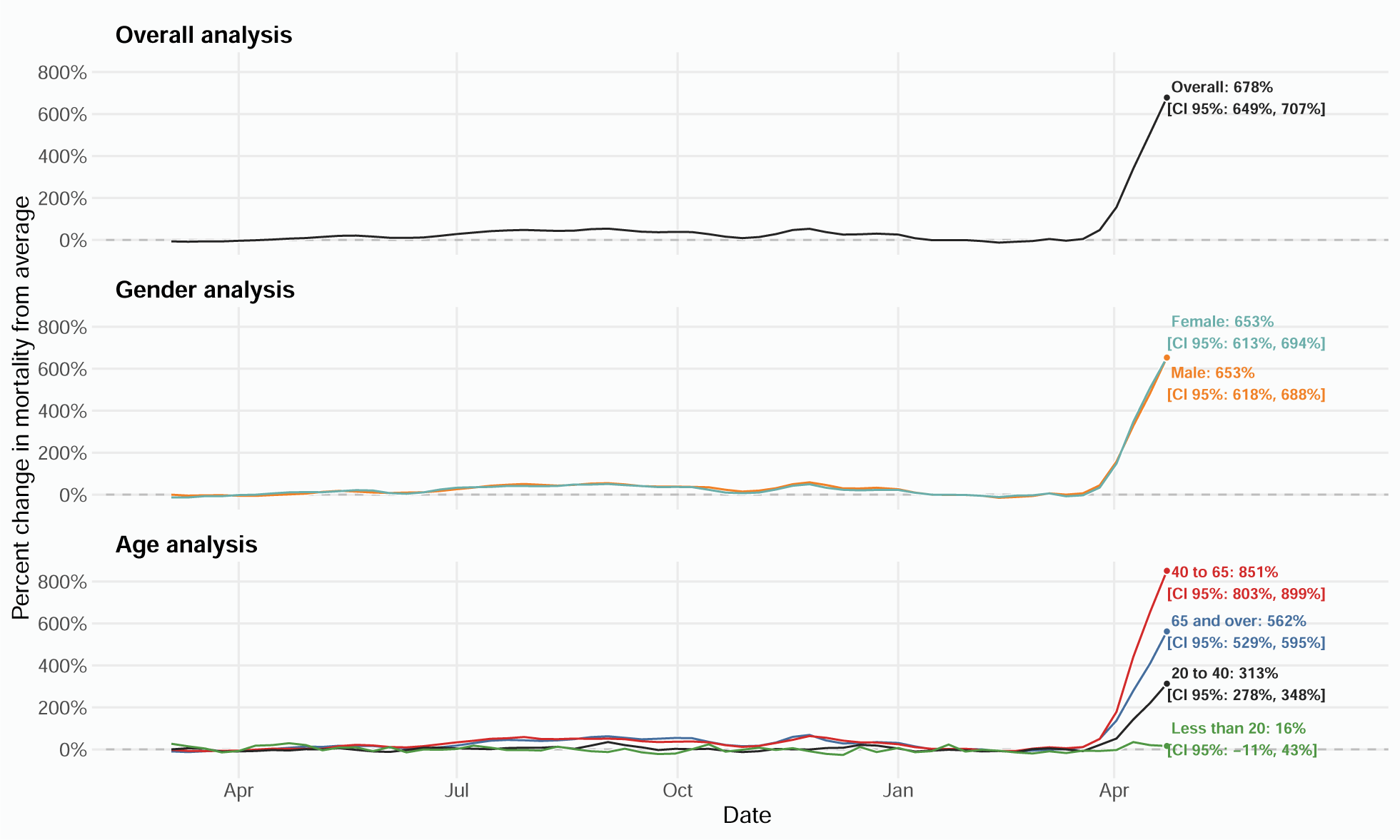
Estimated percent change in mortality from average in Gujarat, India, from March 2020 to April 2021. The solid-curves represent percent changes from average mortality for each group. 95% confidence intervals were omitted for better readability. The point estimate and corresponding 95% confidence intervals for April 16, 2021, the week of peak excess mortality, are displayed in text on the right and highlighted with a data point.

Starting from March 2021, we once again observed an increase in mortality - this time precipitous - that varied by age but not by sex. For this period, we found an overall increase in mortality of 339% [95% CI: 330%, 347%]. For males and females, we estimated increases of 326% [95% CI: 316%, 336%] and 333% [95% CI: 321%, 345%], respectively. For the groups, 20 to 40, the 40 to 65, and the 65 years and over, we found increases of 163% [95% CI: 151%, 174%], 433% [95% CI: 419%, 447%], and 283% [95% CI: 273%, 292%], respectively. In our sample, peak mortality occurred in the last week of available data, where the steep positive slope in Figure 2 suggests that excess mortality very likely kept increasing in the subsequent weeks. In this week, we estimated a 678% [95% CI: 649%, 707%] increase in overall mortality. We further found a 653% [95% CI: 613%, 694%] and a 653% [95% CI: 618%, 688%] increase in mortality for females and males, respectively. Conversely, we found substantial variation across age groups. We found no evidence of inordinate mortality for those 20 years and younger. For the 20 to 40 years age group, we found an increase of 313% [95% CI: 278%, 348%]. Of note, the 40 to 65 years age group experienced excess mortality larger than the 65 and over age group. For the former, we estimated an increase of 851% [95% CI: 803%, 899%] and for the latter an increase of 562% [95% CI: 529%, 595%].

### 3.3 Excess mortality by age and by sex for municipalities

Across all 90 municipalities, the largest increases in mortality occurred in the second wave of the pandemic (Figure 3). In the last week of available data, 85 of the 90 municipalities we studied experienced increases in mortality over 100%. Forty-seven of the 90 municipalities experienced increases in mortality over 500% during the same week. Excess mortality differed by age and by sex at the municipal level (Supplemental Figures 4 5). We found this to be particularly true in larger municipalities, where we appraised the size of each jurisdiction via the estimated intercepts in model 1 (Supplemental Figure 6). We further found that over 80% of the municipalities experienced increases in mortality over 100% in all demographic groups except the two youngest age cohorts. Lastly, in over 50% of the municipalities, males and the 40 to 65 years groups experienced an increase of over 500%.

**Figure 3:**
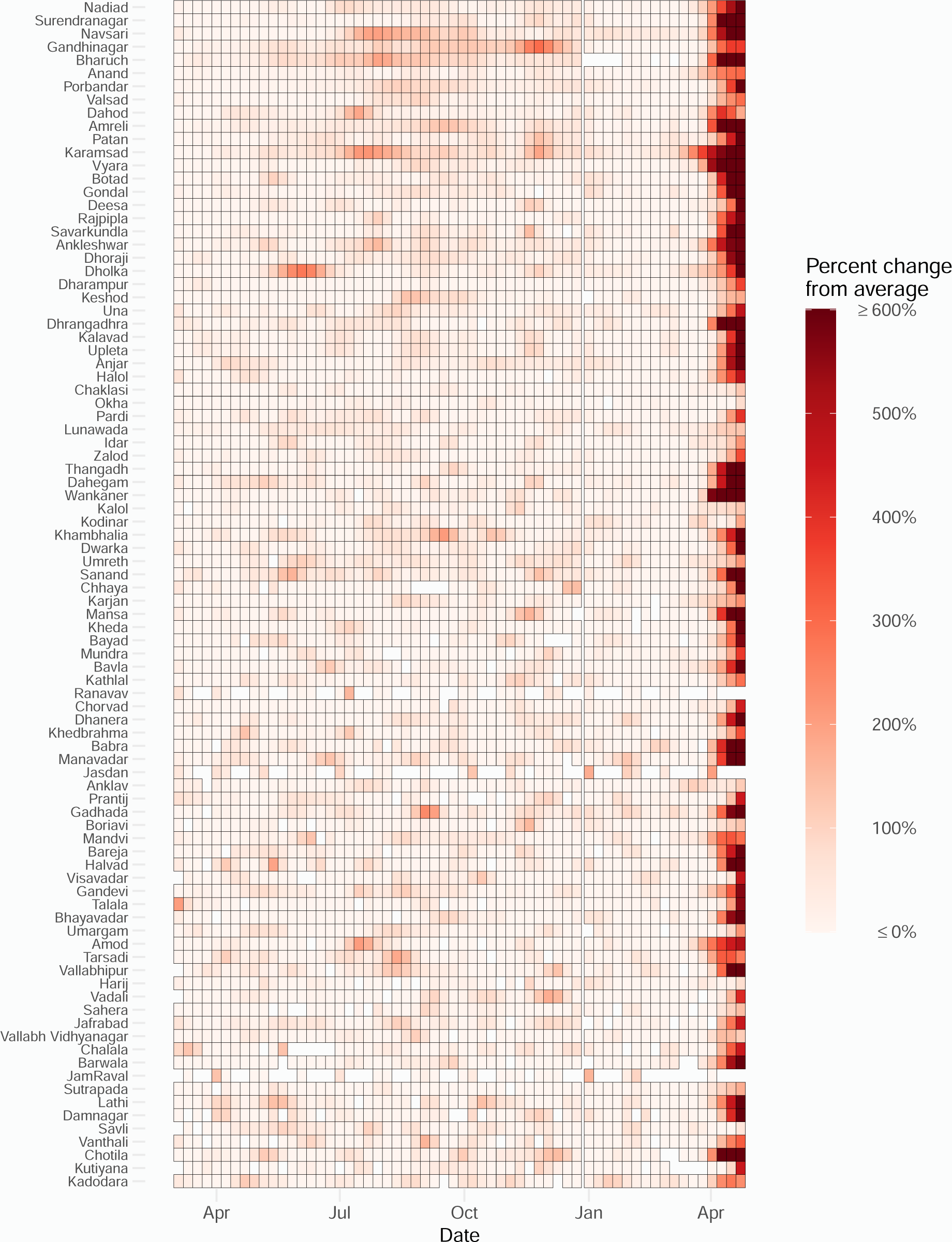
Estimated percent increase in mortality in the 90 municipalities during the Covid-19 pandemic from our marginal analysis.

## 4 Discussion

We describe mortality trends across 90 municipalities in Gujarat, India over the course of the COVID-19 pandemic, until April 2021. The official Covid-19 death count for the entire state of Gujarat is 10,098[30]. However, in a recent hearing to India’s Supreme Court, Gujarat’s government accepted almost 10,000 more Covid-19 deaths[31]. Our results suggest that in these 90 municipalities alone there were 21,300 [95% CI: 20,700, 22,000] excess deaths from March 2020 to April 2021, far exceeding the official count. The 40 to 65 age group experienced the greatest increase from baseline mortality relative to other age groups, and both males and females experienced similar increases in mortality. The vast majority of these excess deaths likely represent direct deaths from COVID-19, in the absence of any other known catastrophe. A small percentage of these would include deaths from the indirect impact of the pandemic, and from causes unrelated to the pandemic.

Our study has several limitations. First, the data only represent around 5% of the population of Gujarat covering 90 of 162 municipalities. These are urban municipalities. With the exception of Gandhinagar, they include neither the municipal corporations of other large urban centers, nor the rural gram panchayats. Though the municipalities were spread across the state, they represent a convenience sample rather than a random sample; we are therefore unable to extrapolate our results to estimate deaths across the entire state. According to the NFHS, a high percentage of deaths are actually recorded in the death registers, rendering our analysis highly representative of mortality in the examined municipalities. The strikingly high mortality is also consistent with media reports and lived experience and likely representative of the general trend across the state. Our results cannot be extrapolated to estimate excess mortality nationally, given limitations with their representativeness and the socioeconomic and health system heterogeneity across India. A study of data from health facilities across several Indian states found that Gujarat had the highest increase in mortality during the pandemic[22].

Second, since the data on the yearly population size for each municipality are not available, for a variety of reasons that include changing jurisdictional boundaries, we were unable to calculate mortality rates and make comparisons across municipalities. For the same reason, we were unable to assess excess mortality rates by demographic indicators. The last published census data are from 2011. While data from electoral rolls are more recent, they do not map to the same geospatial unit of the municipality, and cannot be easily used. We therefore make the assumption that the population remained unchanged between January 2019 and April 2021. Our results will be biased if there was significant migration in or out of the state between our baseline period and the pandemic period. Had population sizes significantly increased (or decreased) over time, our excess mortality estimate would be an overestimate (or underestimate). Because mortality varies strongly with age, both for COVID-19 and all-cause mortality, an ideal comparison would also adjust for age-specific population changes.

Third, we only have baseline (pre-pandemic) data from January 2019 to February 2020. Since the baseline period for fitting the model is relatively short, we may not be sufficiently capturing year-to-year variations in mortality. However, the sharp increase in mortality observed in 2021 is unlikely to fall within the bounds of normal yearly variability in mortality. Finally, there may be lags in recording of deaths in the death registry, and not all deaths may yet be registered. According to media reports, mortality continued to be high, or rose, in May 2021, and is not yet included in the published data or in our estimates.

Despite these limitations, the rapid increase in all-cause mortality, especially during the second wave of the pandemic, is irrefutably high. We estimated a 678% increase [95% CI: 649%, 707%] in deaths in the last week of available data, in April 2021, in the municipalities studied. Our results suggest that April 2021 was the beginning of the second wave, a finding that is in agreement with recent estimates[22, 23]. This increase is among the highest percentage increase in deaths recorded anywhere in the world. In April 2020, Ecuador recorded a 411% increase; in April 2021, Peru recorded a 345% increase[32]. This large discrepancy between official COVID-19 death counts and excess mortality underscores the need to rectify how official death counts are collated. Reliance on death certificates as the single source of truth is sub-optimal, anywhere in the world, when access to health systems, testing availability, and death certification accuracy and completeness are weak.

The high mortality counts across age groups, and especially in the 40-65 years group, warrants further investigation into the impact of underlying social determinants and the efficacy of clinical protocols and public health policies on mortality. The lack of relevant data precludes these necessary analyses. Globally, data on population estimates, testing, and clinical outcomes, where available, have facilitated contextually intelligent public health planning and response. State supported data transparency and availability can in fact help local scientists focus on knowledge generation, and provide citizens and the state the tools needed to strengthen health systems.

## Data Availability

The dataset was made publicly available by The Reporters' Collective here: https://www.wallofgrief.org

## Acknowledgements

We thank The Reporters’ Collective[24] for providing the un-identified data that was also made publicly available via a creative commons license at www.wallofgrief.org

## Author Contribution Statement

RA, BP, AM, and SB conceptualized the paper. BP and SB procured the data and provided local context. RA led the analysis with inputs from AM, MK, SB, CB and RI. RA wrote the first draft, edited by AM and SB, and reviewed by all authors. AM and SB contributed equally.

## Supplementary Material

**Supplementary Figure 1:**
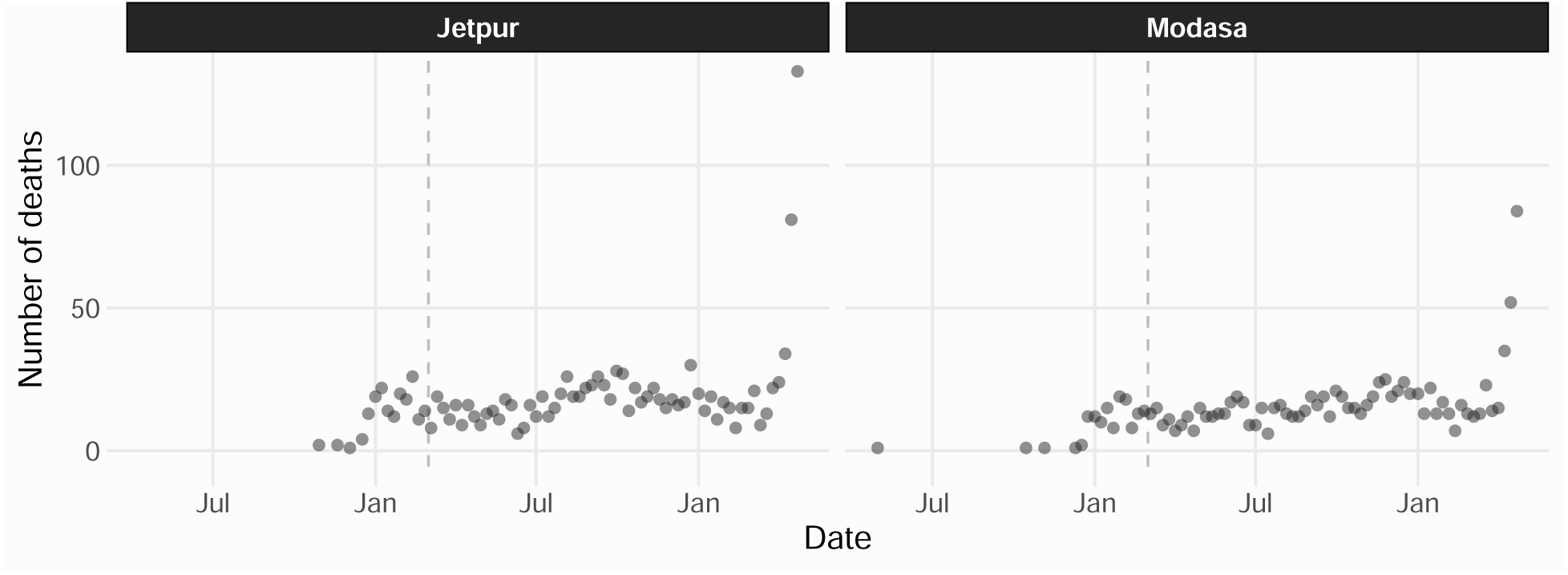
Weekly death counts for Jetpur and Modasa —the two municipalities not used in any of the three analyses.

**Supplementary Figure 2:**
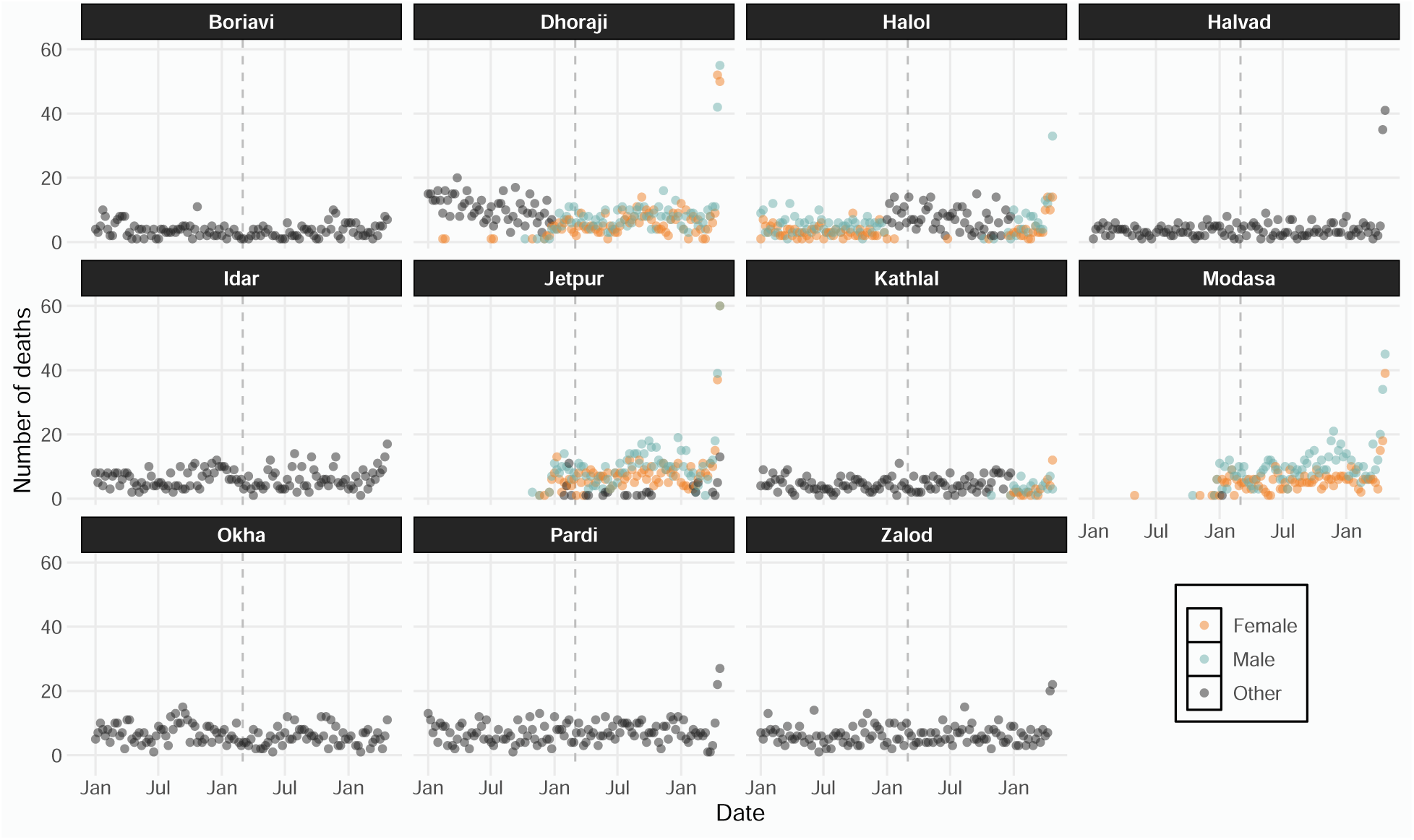
Weekly death counts for municipalities not included in the gender analysis. Jetpur and Modasa are not shown here.

**Supplementary Figure 3:**
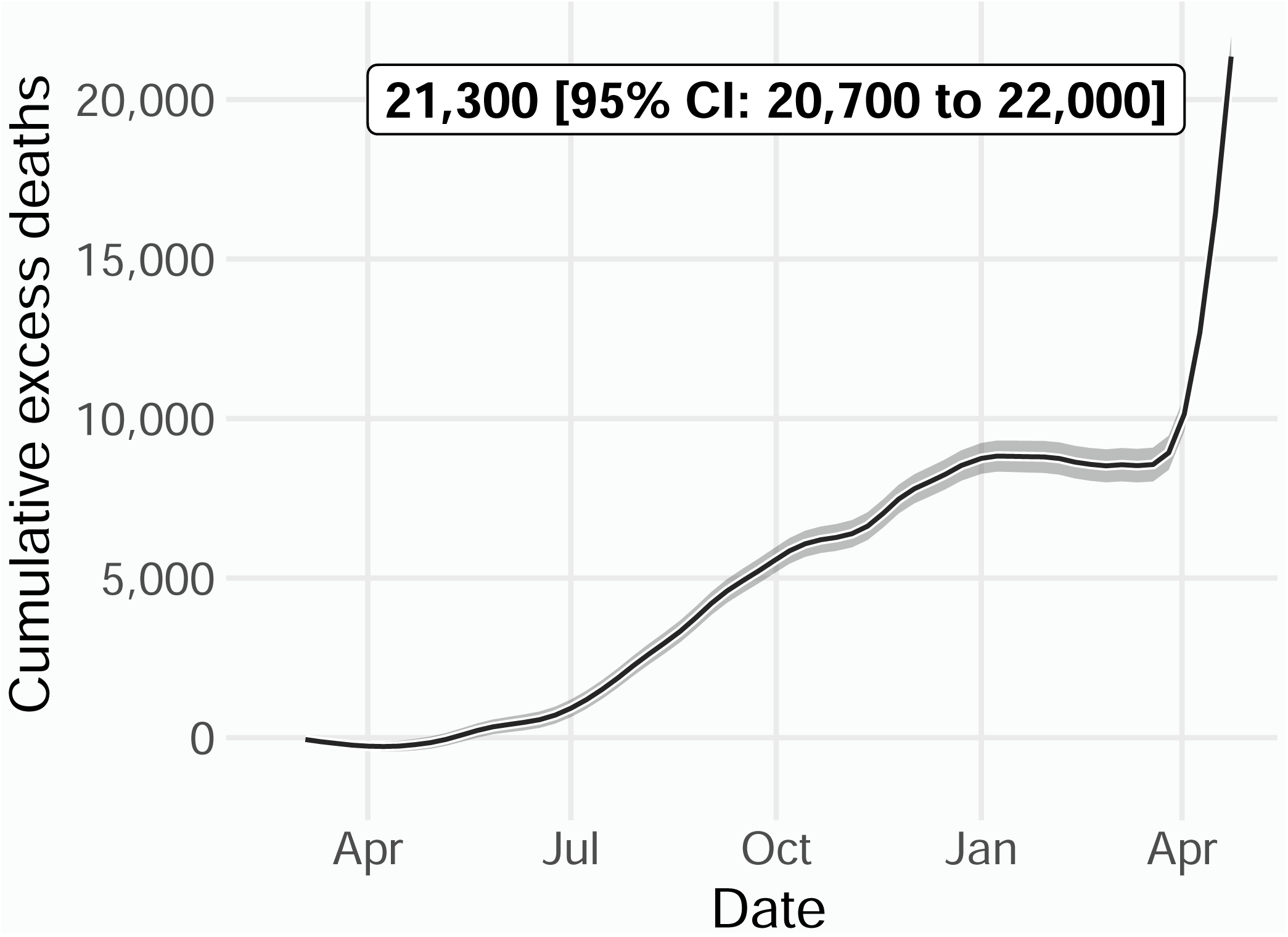
Cumulative excess deaths from March 2020 to April 2021 for all 90 municipalities combined. The cumulative death toll and the corresponding 95% confidence interval are highlighted in text at the top.

**Supplementary Figure 4:**
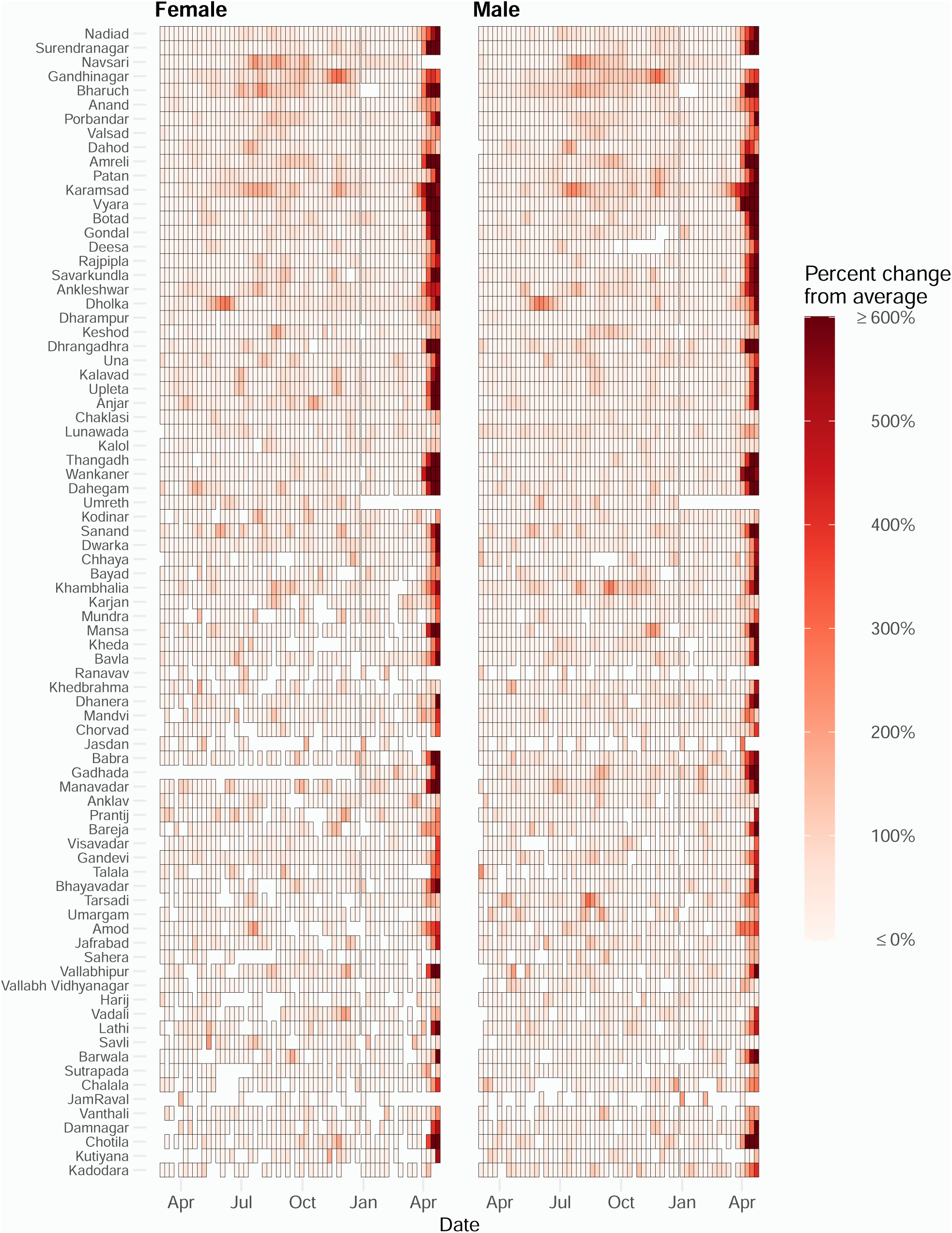
Estimated percent increase in mortality in the 90 municipalities during the Covid-19 pandemic from our gender analysis.

**Supplementary Figure 5:**
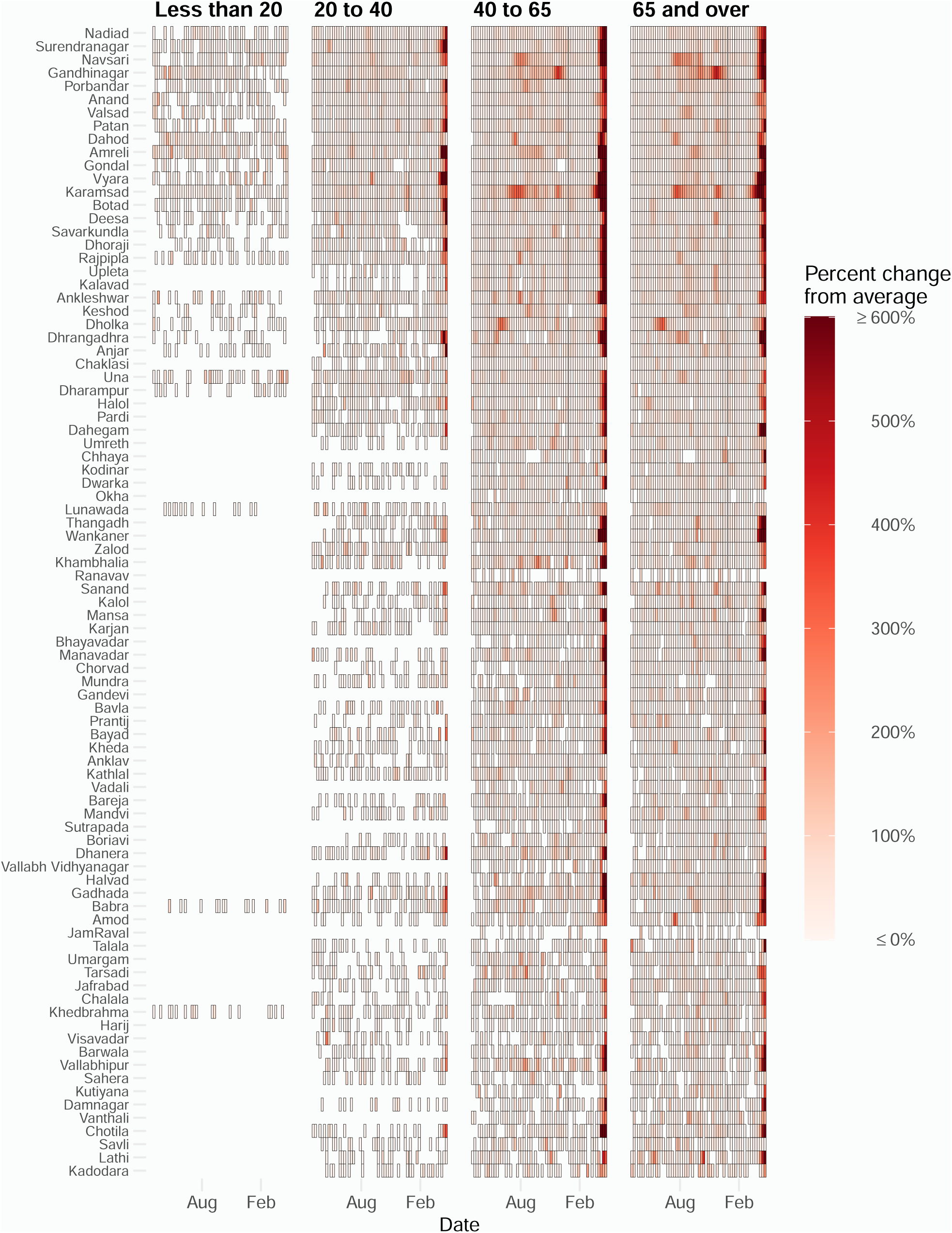
Estimated percent increase in mortality in the 90 municipalities during the Covid-19 pandemic from our age analysis.

**Supplementary Figure 6:**
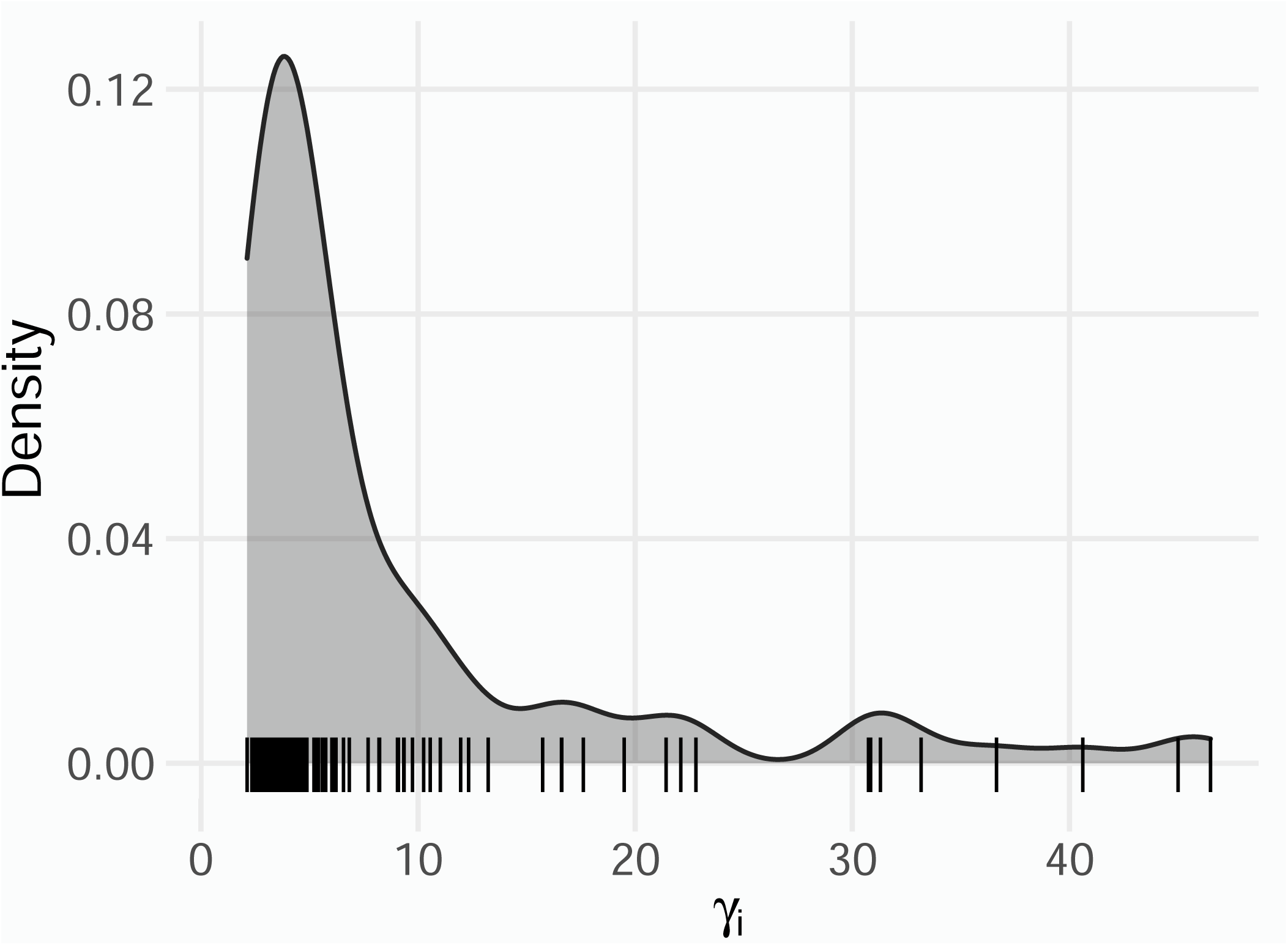
Estimated municipality-specific intercepts from model 1 from our marginal analysis.

**Supplementary Table 1:**
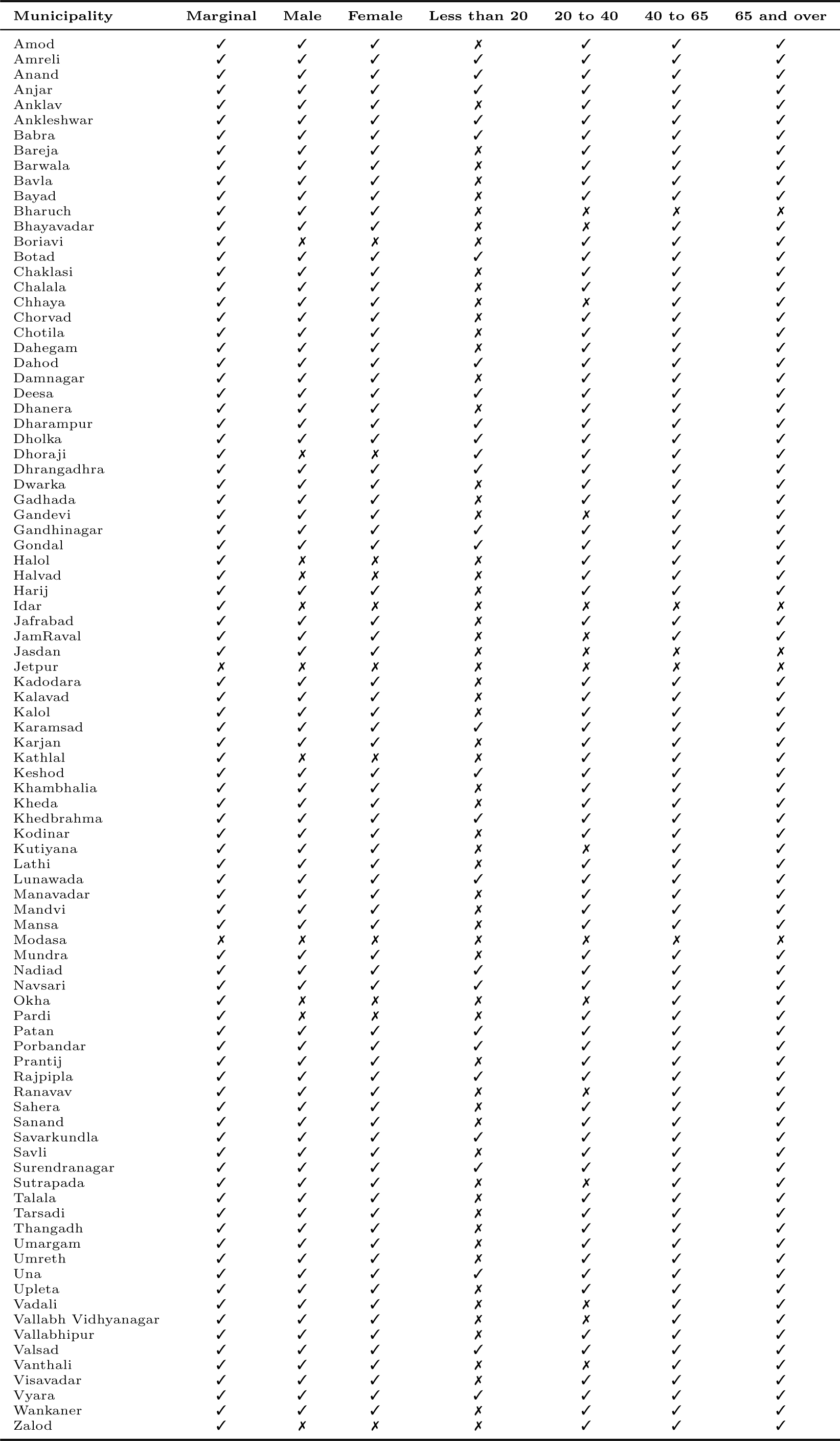
Municipalities included and not included in each analysis.

